# Modeling an epidemic in an imaginary small town

**DOI:** 10.1101/2020.08.31.20185256

**Authors:** Graham Bell

## Abstract

The course of an epidemic in an imaginary small town has been simulated with an agent-based model. The reproduction number *R* of the virus could be counted directly, and was roughly, but not precisely, exponentially distributed. The number of secondary infections was greater for an infection which was itself one of many secondary infections because of environmental heterogeneity, which created variance of *R* among sites and could drive the spread of infection, even when global *R <* 1. Different kinds of intervention were deployed to curtail the spread of infection. Measures applied to the general population, such as closing down sites and services or regulating individual behaviour, did not reduce the total number of individuals infected during the epidemic unless they were maintained until the virus became extinct. This was primarily because measures taken to reduce indirect transmission tended to increase direct transmission, and vice versa. Consequently, the overall effect of any combination of interventions was much less than the sum of their separate effects. On the other hand, the quarantine of infected or exposed individuals was effective in driving the virus to extinction and caused a permanent and substantial reduction in the number of cases.

## INTRODUCTION

Burminster is an imaginary small town with a population of about three thousand whose inhabitants experience an outbreak of infectious disease. The people of the town live in houses or apartments, either singly or as couples who may have children at home. The town has several communal institutions: a hospital, a school that admits children of all ages, a nursery for toddlers, a care home for elderly people and a police station. If they do not work in any of these, people work in factories or shops, keep house, or are retired. ‘Factories’ may be manufacturing companies, warehouses, offices or farms; what they have in common is that they employ workers but do not admit visitors; shops, of course, have both workers and visitors. People travel to work together by public transport. After work they may take their children to a playground or spend the evening with other adults at the cinema or theatre or in a pub.

Once the disease has arrived in the community, anyone may come into contact with infected people and themselves become infected. They may be infected directly, by contact with an infected person, or they may be infected indirectly by contact with surfaces in the home, the workplace or the transit system that have been contaminated by infected people. As the disease continues to spread, the town authorities may decide to take action. If their measures turn out to be effective, the authorities may decide to relax them after a while, but they may be resumed if the disease then begins to flare up again.

The epidemic eventually runs its course and dies out once enough people have been infected and either succumbed or recovered and become immune. A complete record of all the infections has been made during the epidemic. This includes how, where and when each case occurred, as well as the history and genealogy of the pathogen itself. The epidemic is, of course, as imaginary as the town: it is the output of an agent-based simulation. The simulation tracks every person in Burminster during their daily activities, perceives when they are at risk of infection and decides whether or not they become infected. Infection and transmission are stochastic events whose course is decided by rules rather than equations. The program responsible for the simulation, together with more detailed information about its structure and operation, can be downloaded from the author’s website at http://biology.mcgill.ca/facultv/bell/. The age structure and occupations of the inhabitants are close to the average for towns in Britain or Canada, and the case fatality rate of the infection is similar to Covid-19. The program is much too generalized to be used as a predictive tool or a guide to action, however. Its object is rather to investigate the general consequences of interventions during epidemics, and in particular whether the overall effect of a combination of interventions is the sum of their separate effects, and whether any intervention or combination of interventions reduces the total number of people infected during the epidemic.

## METHODS

### Disease transmission

The virus may be transmitted directly from an infected to an uninfected person when they meet at home, at their workplace, when visiting a workplace (for example, a shop), at a playground or pub, or when travelling. The source of each new infection is known, so the number of new infections stemming from each infected person (the ‘reproduction number’ *R* of the virus) can be counted directly. Furthermore, the complete genealogy of the virus metapopulation can be extracted.

If the virus can be transmitted indirectly by touching a contaminated surface its complete genealogy cannot be obtained because the ancestry of the virus responsible is not known with certainty. The average reproduction number can be estimated, because the ratio of direct to indirect transmission is known, but its distribution cannot be obtained by direct count.

A person who has become infected incubates the virus for a certain period of time before displaying symptoms. In some cases these symptoms become severe and the person is hospitalized (if beds are available) and might die. People who recover (the great majority) are immune to further infection for some time, or indefinitely. The detailed time course of the epidemic depends on the values specified for the parameters of the model, for example transmission probabilities, so that an extensive exploration of parameter space is impracticable. Instead, I have chosen a set of parameters which generates an epidemic with reasonable dynamic properties. This approach rules out any attempt to predict the course of any particular epidemic, or the effect of any particular intervention, but makes it possible to investigate the general features of epidemics and the effects of measures to curtail them in a fully transparent system with agents obeying simple rules of behaviour. It is not intended to replace mathematical models such as the covid models described recently by Flaxman et al. (2020), Kissler et al. (2020), Lavielle, Faron & Zeitoun (2020) and Morozova, Li & Crawford (2020). Rather, it supplements them by showing how a different modelling approach leads to broadly similar conclusions.

### Interventions

As the epidemic progresses, the town authorities either let it take its course or decide to intervene in an attempt to slow the spread of the disease. They may decide to close (1) the nursery and school, (2) the non-essential workplaces and shops, or (3) the playgrounds and pubs. They may decide (4) to curtail public transport or (5) forbid visits to patients in the hospital or to elderly residents of the care home. These five options for shutting down sites or services might be combined in any way. The town authorities may also advise or enforce restrictions on behaviour, such as (1) social distancing (which reduces the number of encounters with other people); (2) the wearing of masks (which reduces the probability of direct infection per encounter); (3) the wearing of gloves (which reduces the number of contaminated objects touched); or (4) frequent handwashing (which reduces the probability of infection from touching a contaminated object). They may also (5) organize the regular cleaning of public spaces to reduce the length of time that the virus persists on contaminated surfaces. These five kinds of behavioural modification might also be combined in any way. People showing symptoms of disease may also be placed in quarantine, which can be extended to family members or to the people they work with. The authorities may put in place none or any or all of these measures or any combination of them. The effectiveness of any intervention may well depend on compliance, however, so on any given day each person can decide (with given probability) whether or not to comply with the measures that have been put in place by the town authorities.

Whatever pattern of intervention is decided on, it must be imposed at some point, depending on the progress of the epidemic. In Burminster, the authorities act after the number of new cases has risen consistently in recent days, but any other criterion might be used instead. Once any intervention has been put in place, it must be lifted sooner or later, and this decision, like the decision to impose it in the first place, will depend on the course of the epidemic. The measures taken by the authorities in Burminster are relaxed when the number of current cases has been below its peak value for a specified period of time, but again other criteria might be used. If the disease flares up again, the measures can be imposed again. It would be unrealistic to suppose that the authorities can react instantly to changes in disease incidence, of course, so restrictions are imposed or relaxed only after the criterion adopted has been satisfied for a specified period of time. I investigated in detail a situation in which the authorities impose restrictions after the number of new cases has risen every day for the past week, and relax these restrictions when the number of current cases has been less than its peak value every day for the past two weeks.

## RESULTS

### The virus metapopulation

The reproduction number. The epidemic begins when a single individual, chosen at random, becomes infected. The number of individuals who are subsequently infected by this initial host is the basic reproduction number *R_0_*. The one conclusion from theoretical epidemiology that everyone knows is that an epidemic will spread if *R_0_* > 1 (Anderson & May 1992). *R_0_* is a theoretical concept, however; it is difficult to estimate directly in real populations and is usually inferred from rates of spread, or set as a parameter in theoretical models (Li, Blakely & Smith 2007; Breban, Vardavas & Blower 2007). The beginning of an epidemic is inevitably dominated by stochastic events, because, regardless of the size of the population, the number of infected individuals is initially low. In real populations or agent-based models the virus may fail to spread (Rahmandad & Sterman 2008) and instead becomes extinct (with probability 1/*R_0_*; Chiang 1980). The necessary condition for an epidemic to be initiated is that the founding virus infects at least one new host and, should it infect only a single new host, that its lineage includes at least one virus that infects at least two new hosts. The process is analogous to the initial spread of a beneficial allele under natural selection (see Crow & Kimura 1970 p 418; Gerrish & Lenski 1998).

In agent-based models, the reproduction number can be counted directly from the output (Keeling & Grenfell 2000; Green, Kiss & Kao RR 2006). The closest approach to *R_0_* is the average reproduction number early in a developing epidemic, which I shall call the historical reproduction number *R_H_*. It is calculated over all past infections that have been cleared by recovery or death, but does not include current infections, since some of these will eventually give rise to secondary infections but have not yet done so. The value of *R_H_* at any particular time corresponds to the *R_t_* of previous authors such as Flaxman et al. (2020).

In a small town such as Burminster the epidemic proceeds in two phases. The first phase is largely stochastic: a few secondary infections are generated, driving *R_H_* to high values which fluctuate irregularly from day to day. There is no rapid and sustained rise in cases because, although susceptible individuals are very frequent, infected individuals are very rare, and consequently *R_H_* often drops soon afterwards to values close to 1. If the fuse has been successfully lit, however, the epidemic now enters a second phase in which deterministic forces predominate after a sufficient number of hosts have been infected: *R_H_* increases until limited by the supply of susceptible hosts, after which it declines asymptotically towards *R_H_* → 1 in the later stages of the epidemic (Figure 1).

**Figure 1.**
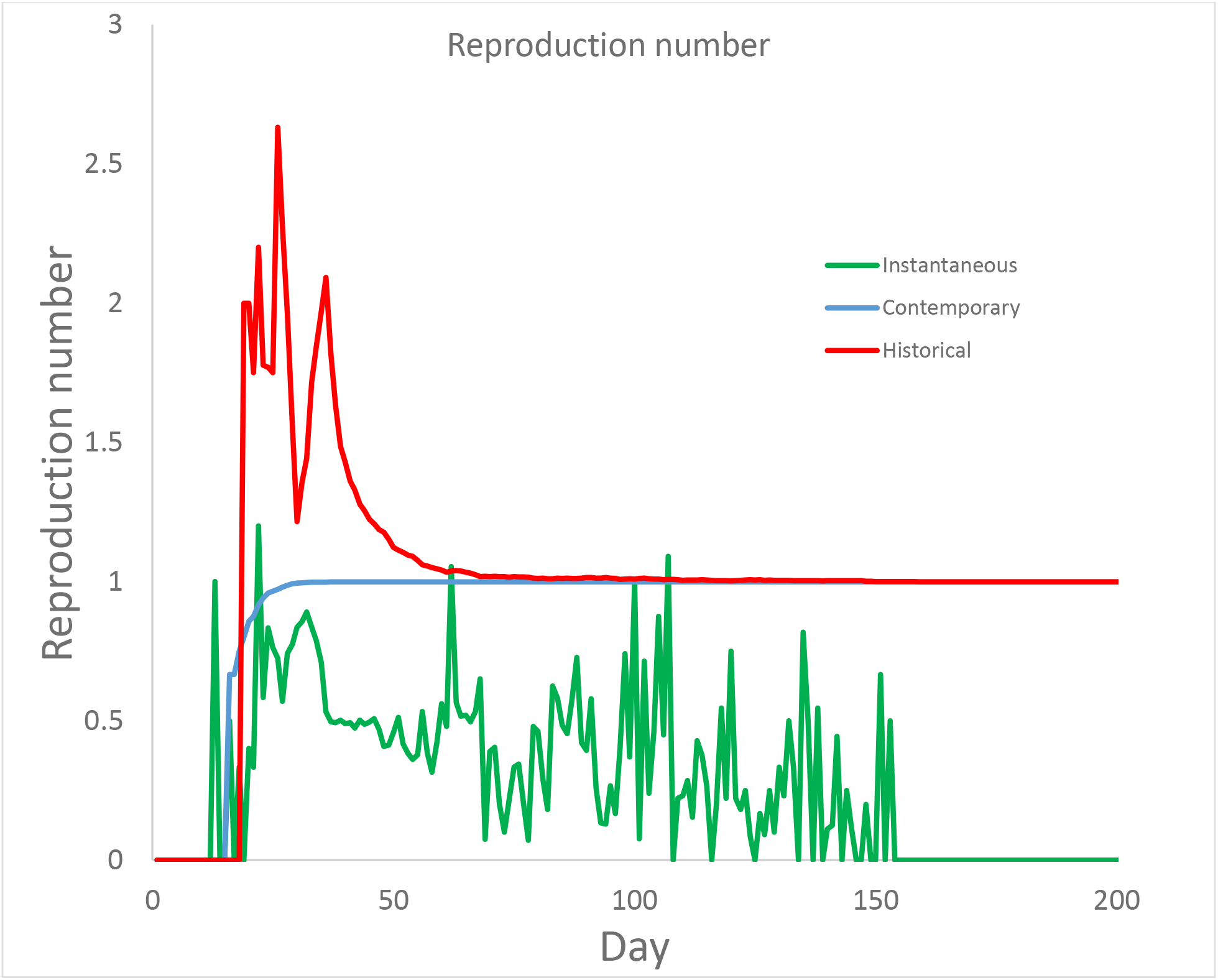
The reproduction number over the course of the epidemic. Green line: instantaneous reproduction number R_I_. Blue line: contemporary reproduction number R_C_. Red line: historical reproduction number R_H_.

Other ways of calculating an average reproduction number can be informative. The contemporary reproduction number *R_C_* is the mean reproduction number of all virus populations (that is, infected hosts) that have lived till now, including those currently alive. This is a well-behaved statistic that rises smoothly over time from zero at the beginning of the epidemic, when the ancestral propagule has yet to be transmitted, to an asymptote at *R_C_* → 1 at the end, when *R_C_* = *R_H_*. The instantaneous reproduction number *R_I_* is the probability that a current infection will give rise to a secondary infection in the next short interval of time. This is the measure that drives change in the number of daily cases, which will increase if *R_I_* > 0.5 (neglecting the possibility of multiple secondary infections and assuming that the current infection has not yet been cleared). The probability that an epidemic will result is far less stringent, however. The probability that a new infection will eventually give rise to more than one secondary infection is the binomial expansion of *[R_I_ +* (*1-R_I_*))*^t^*, where *t* is the infective period, neglecting the first two terms, i.e. 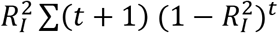, which can be substantial even when *R_I_* is small.

If either the virus or the host population is heterogeneous then the criterion *R_0_* > 1 may not apply, even in theory. If there is more than one strain of the virus then the strain which spreads most rapidly will tend to supplant others, and may cause an epidemic even if it is initially rare and all other strains have *R_0_* < 1. This is an example of evolutionary rescue (Gomulkiewicz & Holt 1995; Bell & Gonzalez 2011; reviewed by Bell 2017). The criterion may also fail if the environment is heterogeneous: if the host inhabits several regions, then an epidemic can be sustained by the movement of susceptible hosts from one region to another, where without movement it would die out (Cross et al. 2007; Smith, Gordon & Heffernan 2009). Burminster is a single region, but it is divided into compartments where different sets of individuals work, play or travel, and the epidemic often spreads rapidly in a particular workplace, care home or travel route before taking hold in the general population. These instances show that the reproduction number, however defined, is best interpreted as the property of a lineage rather than of the population as a whole.

Reproductive value. The lineage of virus responsible for any given infection may give rise to 0, 1, 2, … *x* descendent lineages, each representing the infection of a new host. I shall refer to the value of *x* as the rank of an infection. All lineages will eventually die out when the epidemic comes to an end and the virus is locally extinct. At this point, the frequency of ancestors which left *x* descendent lineages is *f*(*x*), assessed over all the virus populations that existed during the epidemic. The distribution of *f*(*x*) is roughly, but not exactly, negative exponential (Figure 2).

**Figure 2.**
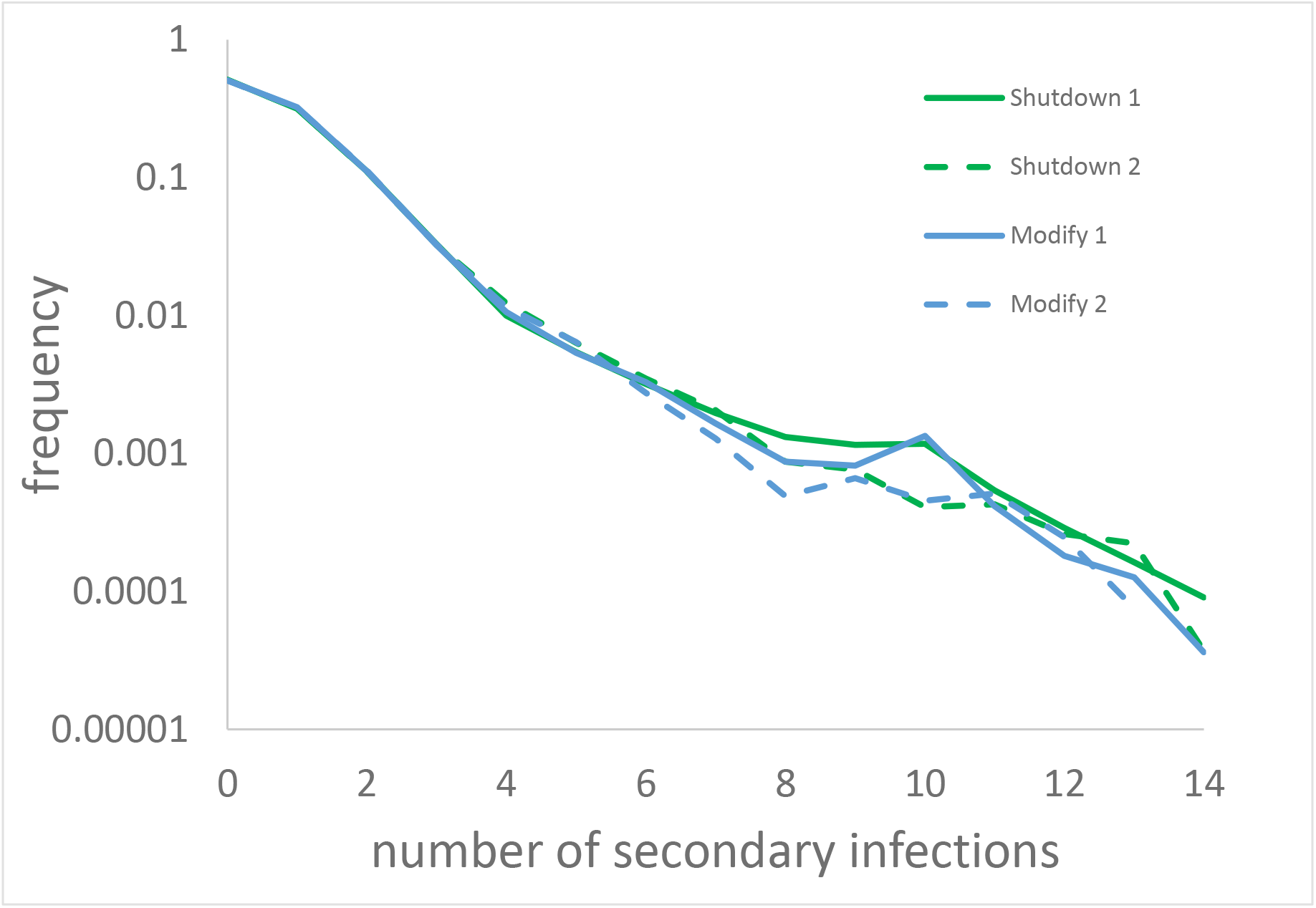
Frequency distribution of the number of secondary infections stemming from a primary infection by direct transmission. Two replicates of all 32 possible combinations of five shutdown interventions and of five behaviour modification interventions. Replicates have the same parameter values but different random number seeds.

It departs from a strictly exponential distribution because the frequency of infections which fail to give rise to any secondary infections is somewhat less than would be expected, whereas the number which give rise to many infections is greater. The shape of the distribution is not sensitive to the type of intervention, nor even to whether any intervention is attempted.

Two secondary statistics can be derived from this raw frequency distribution. The first is simply *x f*(*x*), giving the average number of new infections stemming from infections of given rank. This is modal at *x* = 1, and this class contributes about 40% of new infections; about two-thirds of all new infections are contributed by *x* = 1 or 2. The epidemic is thus driven primarily by infected hosts who infect one or two others. We can go further to calculate the expected number of future infections stemming from an infection of given current rank. The probability that the ancestral lineage will itself give rise to *x* descendants is 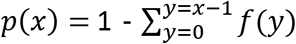; likewise the probability that a lineage that has already produced *x* descendants will survive to produce *y* descendants (*y>x*) is 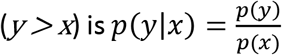. Since a single descendant lineage is produced for each unit increase in rank, the expected number of future descendants that will be produced by a virus of current rank *x* is 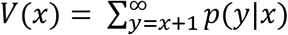. This is similar to the concept of reproductive value in the theory of age-structured populations (Barton & Etheridge 2011). The value of *V*(*x*) is a constant independent of rank for a purely exponential distribution, but the actual distribution of *f*(*x*) leads to *V*(*x*) being modal for rather large values of *x* (*x*= 6 or 7). That is, the number of secondary infections tends to be greater for an infection which was itself one of many secondary infections. The unexpectedly repeatable property of infectivity is not attributable to genetic variation (the virus is assumed to be genetically uniform) but rather comes about because an individual living or working in conditions that are especially conducive to virus transmission is likely to have been infected by a host who has already infected many others, and is in turn itself likely to infect many others.

## The host population

### General dynamics

The general dynamics of infection and intervention are illustrated in Figure 3. When no intervention is attempted, the epidemic follows a familiar pattern. The number of cases rises rapidly to a peak and then slowly declines, ending after about 200 days when about 50-60% of the population has become infected and either died or (the great majority) recovered. Any intervention or combination of interventions reduces the peak and causes a drop in the number of infections. When the intervention is relaxed, the number of current infections rises to a new, lower, peak and afterwards declines as the epidemic ebbs. If the intervention is not relaxed at all during the course of the epidemic, there is no ‘second wave’ and the number of current infections quickly declines to zero.

**Figure 3.**
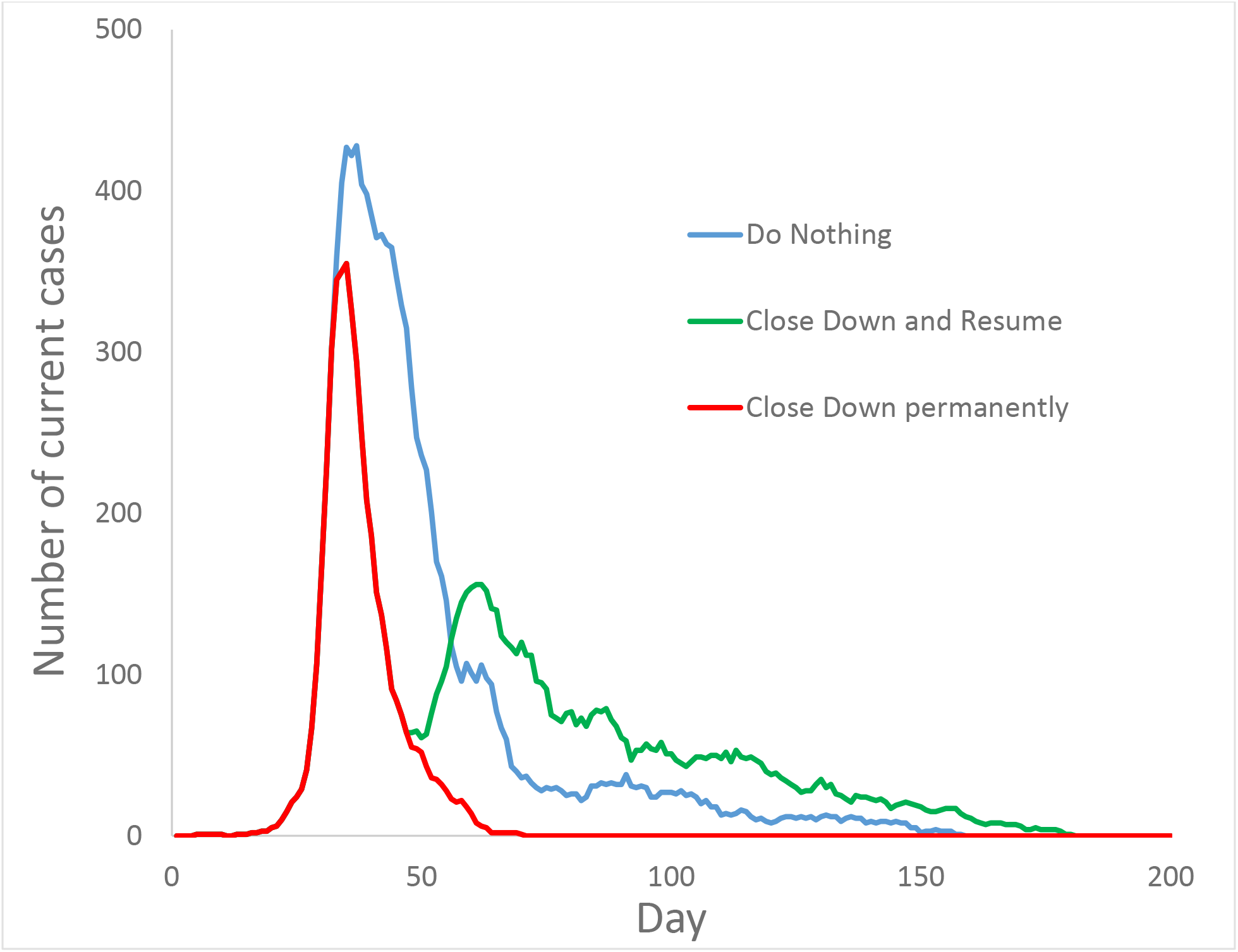
General dynamics of infection and intervention. Blue line: do nothing. Green line: close down all non-essential sites, but resume if current cases have been less than peak for at least 14 consecutive days. Red line: close down all non-essential sites for the duration of the epidemic.

### Interventions

Two kinds of action might be contemplated by the authorities. The first is an intervention applied to the general population. It may involve closing down sites (such as schools or shops) or services (such as public transport). It might also attempt to modify behaviour, such as requiring masks to be worn in public spaces. The second kind of action is applied only to infected individuals, and amounts to the quarantine of more or less inclusive groups. I shall describe first the effect of interventions applied to the general population, before describing briefly the outcome of quarantine.

Any intervention has both general and specific effects. The specific effects follow from the type of intervention. For example, social distancing reduces the number of encounters, while wearing a mask reduces the transmission rate per encounter. Both are intended to reduce the frequency of direct transmission, and this is reflected in the outcome; this merely confirms that the model works as intended.

The general effect is the overall reduction in the number of people who have become infected during the course of the epidemic. There is no systematic tendency for any intervention or combination of interventions to cause an overall reduction in the number of infections over the course of the epidemic, provided that the intervention is relaxed before the virus is extinct (because *R_H_* has fallen below 1 for a sufficiently long period of time) and the epidemic ends. On occasion, a particular combination may be associated with fewer total infections; but this outcome is generally not repeated when the model is run again with identical parameter values but a different random number seed. Consequently, analysis of variance shows no effect of the particular kind of intervention. The reason for this surprising and rather disappointing conclusion is an interaction between modes of transmission that I shall call ‘compensation’.

### Compensation

Behavioural regulations target either direct or indirect transmission: social distancing and wearing masks is intended to reduce direct transmission from exhaled droplets, whereas wearing gloves, washing hands or cleaning public spaces are intended to reduce indirect transmission from touching contaminated surfaces. These interventions succeed in their objective of reducing either direct or indirect transmission. They also have an unintended consequence, however: those that reduce direct transmission tend to increase indirect transmission, while those that reduce indirect transmission tend to increase direct transmission. When one transmission route is blocked, there is a compensatory expansion of the other (Figure 4). The extent of this compensation is such that the overall reduction in cases is only modest atbest. Hence when all combinations of interventions are considered there is little if any average effect on the eventual number of cases.

**Figure 3.**
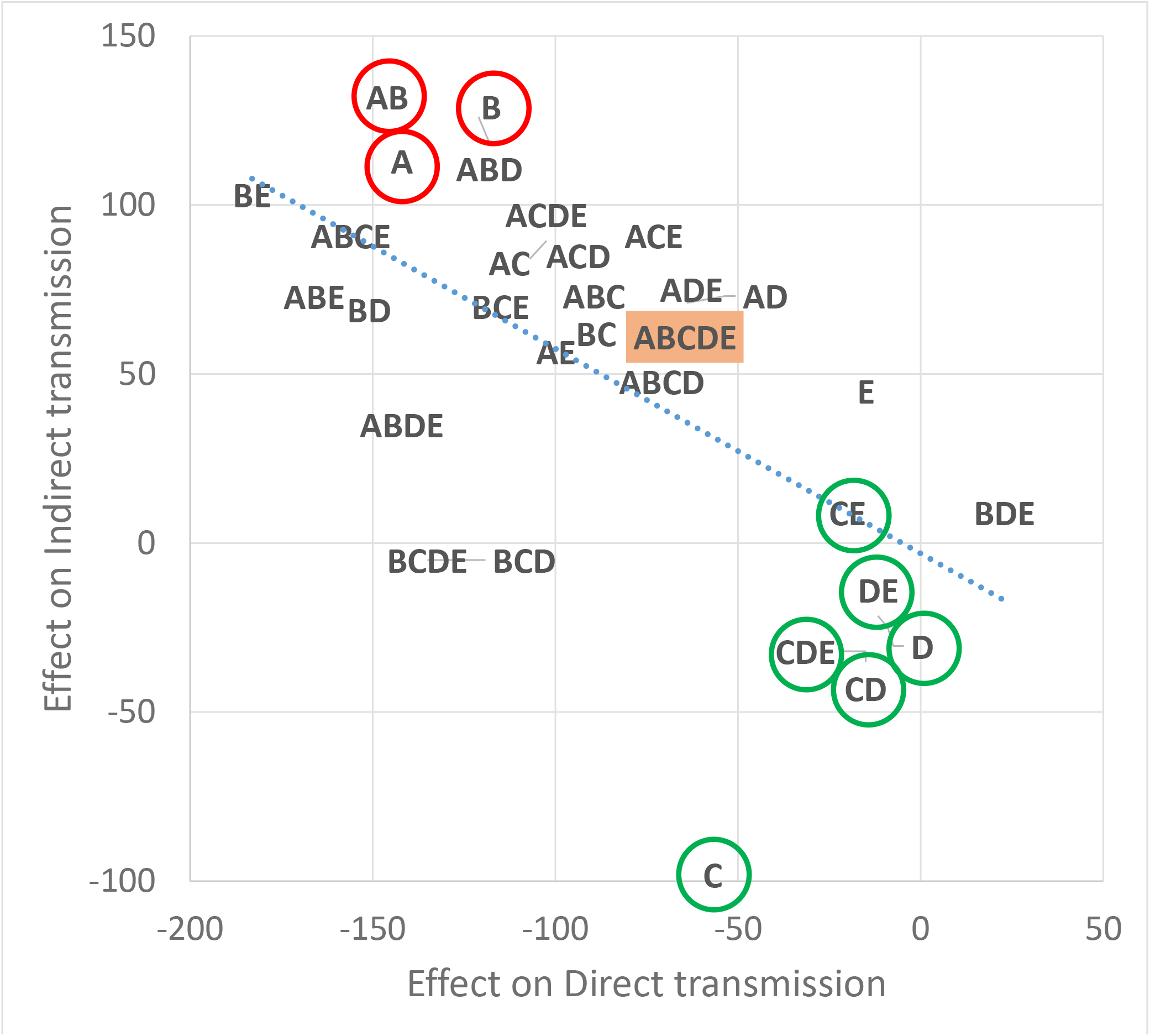
Compensation. The relation between direct and indirect transmission, calculated as the difference in the total number of cases between a combination of interventions and no intervention. The interventions are behavioural modifications: A social distancing (which reduces the number of encounters with other people); B the wearing of masks (which reduces the probability of direct infection per encounter); C the wearing of gloves (which reduces the number of contaminated objects touched); D frequent handwashing (which reduces the probability of infection from touching a contaminated object); E organize the regular cleaning of public spaces to reduce the length of time that the virus persists on contaminated surfaces. Red circles: A and B interventions (reduce direct transmission). Green circles: C,D and E interventions (reduce indirect transmission). The combination of all interventions ABCDE is shaded.

The underlying cause of compensation is that any intervention which successfully blocks either direct or indirect transmission necessarily increases the number of susceptible hosts, relative to direct and indirect transmission. These hosts are then vulnerable to infection by the other transmission route. Provided that the virus can be propagated through either route, the overall number of cases will not be substantially reduced by blocking one of them. Conversely, if indirect transmission does not occur then interventions that hinder direct transmission cause a permanent reduction in the number of cases.

Shutting down sites or services will block both direct and indirect transmission. It will inevitably block one more effectively than the other, however, and compensation will still occur. This also applies to combinations of behavioural modifications that affect both direct and indirect transmission, such as social distancing and frequent handwashing.

Compensation can be viewed as a special case of the general non-additivity of the effects of interventions that are simultaneously applied. If the effect of intervention A is α and the effect of intervention B is β, then the combination A&B will have a predicted effect α+β if effects are additive. The output from the model, however, shows no correlation between the observed and predicted effects of pairwise combinations of interventions.

### Quarantine

The quarantine of symptomatic individuals, rigorously enforced, precludes both direct and indirect transmission. This reduces the reproduction number of the virus and accelerates its extinction, thereby leading to a permanent and substantial reduction in the total number of cases during the epidemic. Its effect does not seem to be very sensitive to compliance; 5% delinquency does not greatly impair its efficacy, much of which is retained even if 30% of symptomatic individuals ignore the regulation. Moreover, its effect scales with effort: quarantining all family members is more efficacious than quarantining only single individuals, and quarantining all workplace colleagues is more effective still. The effectiveness of quarantine stems from the assumption that it can be maintained in force indefinitely because it is applied only to infected individuals, rather than to the population at large. Provided that compliance is sufficient to reduce the reproduction number of the virus below replacement, quarantine will drive the virus to extinction and thereby end the epidemic with fewer cases.

### Vaccination and immunity

The situations that I have described all assume that infected individuals enjoy complete permanent immunity when they recover. If a vaccine is administered the epidemic is halted immediately because there are too few susceptible individuals to sustain it. If individuals who have recovered or who have been vaccinated are immune only for a limited period of time, however, the disease becomes endemic. If the period of immunity is short, susceptible individuals will be numerous and a sustained high number of current cases persists indefinitely. As the period increases, the pool of susceptible individuals shrinks and the current number of infections declines. This creates a time lag between the initial peak phase of the epidemic and the loss of immunity of the individuals who were then infected. If the virus has not become completely extinct, this time lag leads to a new outbreak, and subsequently to a recurrent epidemic. As before, interventions such as shutting down sites and services have little effect on the long-term total number of cases, whereas quarantine reduces the number substantially even when immunity is short-lived.

## DISCUSSION

The main conclusion of these simulations is that no intervention or combination of interventions applied to the population at large will substantially reduce the total number of infections over the course of the epidemic, unless the intervention is sufficiently severe and prolonged to drive the virus to extinction. In Burminster, the threshold for extinction is about 10 weeks when all non-essential sites are shut down. In other communities, however, the threshold will depend on the value of parameters such as transmission rates, the effect of the intervention on these parameters, and the size of the population. This seems to be consistent with the conclusion that measures such as social distancing may have to be maintained for months in order to block the covid epidemic (Lai et al. 2020). Quarantine is more effective simply because it can readily be maintained in force for long enough to drive the virus to extinction. Once the virus has become locally extinct the epidemic comes to an end, and new infections can appear only through immigration.

A year after the epidemic had begun, the Burminster authorities might be imagined to hold a meeting to discuss the consequences of the measures they had taken. With the benefit – denied to any real administration – of being able to evaluate the success of any particular strategy, they would find that shutting down sites and services or enforcing modes of behaviour enabled them to modulate the dynamics of the epidemic and to channel the pattern of infection, without having much effect on the total number of cases, unless these interventions remained in force for a long time. Imposing a quarantine, on the other hand, damped down the epidemic and reduced the number of cases substantially. No doubt they would hope for permanent immunity against re-infection and the timely release of an effective vaccine. In the meantime, they would have to plan for the next epidemic in the light of their experience over the past year.

## Data Availability

The code and output of the model are available on the author's website at https://mcgill.ca/biology/graham-bell or by request to the author at.

## Acknowledgments

This work was funded by a Discovery Grant from the Natural Sciences and Engineering Research Council of Canada. I am grateful to Thomas Bell, Austin Burt, Vassiliki Koufopanou and Michael Rosinger for comments on an earlier version.

## Notes

### Competing Interest Statement

The authors have declared no competing interest.

### Clinical Protocols

https://mcgill.ca/biology/graham-bell

### Funding Statement

This work was funded by a Discovery Grant from the Natural Sciences and Engineering Research Board of Canada.

### Author Declarations

No approvals or exemptions for this work are necessary.

